# Initial Treatment of Glaucoma with Selective Laser Trabeculoplasty: Economic Impact from the Perspective of the Brazilian Public Health System

**DOI:** 10.1101/2024.07.21.24310769

**Authors:** Ivan M. Tavares, Flavio E. Hirai, Diogo F. C. Landim, Paola Zucchi

**Affiliations:** Department of Ophthalmology and Visual Sciences, and the Discipline of Health Management and Economics, Paulista School of Medicine, Universidade Federal de São Paulo; Master’s degree program in Health Economics and Management, Faculdade Paulista de Ciências da Saúde, Hospital São Paulo, SPDM. São Paulo, Brazil

**Keywords:** Glaucoma, Trabeculoplasty, Laser Therapy, Cost Analysis, Health Care Cost

## Abstract

**Objective:** To evaluate the economic impact on the Brazilian Public Health System (SUS) of the initial treatment of glaucoma in two scenarios: (1) the traditional continuous clinical treatment with hypotensive eye drops and (2) the single treatment with Selective Laser Trabeculoplasty.

**Methods:** Economic impact analysis was conducted in three scenarios, from the least to the most conservative, for the two initial treatment methods for open-angle glaucoma in a hypothetical cohort of 5,000 individuals. Projections were then made based on a prevalence of 3% of the Brazilian population in 2021.

**Results:** All three scenarios analyzed showed a significantly lower economic impact for Selective Laser Trabeculoplasty on the Brazilian Public Health System over one to five years, with a favorable difference of more than 8 billion US dollars over five years when considering 3% of the Brazilian population over 40 years old in 2021.

**Conclusion:** The economic impact on the Brazilian Public Health System was lower for Selective Laser Trabeculoplasty compared to the use of latanoprost and timolol maleate eye drops as the initial treatment for primary open-angle glaucoma in all scenarios studied over one and five-year periods.

## 1.0 Introduction

Glaucoma is an optic neuropathy characterized by the slow and progressive degeneration of the retinal ganglion cells (RGCs), resulting in typical structural changes of the optic nerve head and retinal nerve fiber layer (RNFL) and corresponding defects in the visual field. RGCs are central nervous system neurons with cell bodies and axons located in the inner retina. The axons organize themselves internally, forming bundles that constitute the RNFL, converge at the optic disc, and form the optic nerve.(1)

It is the second leading cause of blindness worldwide and the leading cause of irreversible blindness, according to the World Health Organization.(2) Similar data were found in the greater São Paulo area(3). Considering only Primary Open-Angle Glaucoma (POAG), in a Brazilian study, its prevalence in people over 40 years old was 2.4% among whites and 3.8% among non-whites(4), increasing with age in all studied populations(1).

Clinical treatment classically begins with hypotensive eye drops, usually from the prostaglandin analog class or beta-blocker, with other medications possibly added for adequate intraocular pressure (IOP) reduction. Each group of hypotensive ocular drug has a primary mechanism of action, such as reducing aqueous humor production, increasing the uveoscleral or the trabecular outflow. Long-term treatment with eye drops often involves side effects and requires good patient adherence.(5,6)

The introduction of laser (light amplification by stimulated emission of radiation) in glaucoma treatment in the latter half of the 20th century significantly impacted the management of various clinical conditions. In glaucoma, the most exploited effects are thermal, mainly with argon laser, and ionizing, with Nd:YAG (neodymium-doped yttrium aluminum garnet) and diode lasers.

Patients with open-angle glaucoma can have their intraocular pressure (IOP) reduced through laser trabeculoplasty. Selective Laser Trabeculoplasty (SLT) involves applying a frequency-doubled Nd:YAG laser directly to the trabecular meshwork. The procedure leads to trabecular remodeling, increasing drainage through this route and thus reducing IOP.(7,8)

Initial glaucoma treatment with trabeculoplasty, despite its temporary effect of around 36 months, positively impacts the patient’s quality of life, reducing the need for eye drops to control IOP and the occurrence of side effects. It is an outpatient procedure with good safety profile, and it can be repeated if needed. In the LiGHT Trial, a multicenter randomized clinical trial comparing SLT with prostaglandin analog hypotensive eye drops, including approximately 350 participants in each group, mostly with early glaucoma, the authors showed that initial laser treatment, compared to eye drops, was safe, more cost-effective, equally or more effective, and less dependent on patient adherence to treatment. On average, in the group treated with trabeculoplasty, 78% of patients had IOP control at the end of 36 months, with 77% of these requiring only one laser treatment session. (9)

Given budgetary constraints, the need for rational use of public resources, and adherence to best clinical practices, the cost of treatments offered by the Brazilian Public Health System (SUS) must be continuously evaluated. In Brazil, studies about costs of glaucoma treatment are scarce. The study by Guedes et al. evaluated the cost-effectiveness of clinical treatment (with eye drops) or laser (unspecified trabeculoplasty) against observation, which is no longer accepted upon diagnosis of the disease. They concluded that both strategies were cost-effective, with a slight advantage for laser treatment, and that both alternatives provided significant quality-of-life gains.(10)

Determining the initial treatment for glaucoma with the least economic impact is imperative for SUS. Although SLT requires specific equipment and a qualified ophthalmologist to be performed, we hypothesize that the economic impact of initial glaucoma treatment using SLT is lower than the traditional use of hypotensive eye drops. Therefore, the objective of this study is to evaluate the economic impact on the SUS of initial glaucoma treatment in two scenarios: (1) the traditional continuous clinical treatment using hypotensive eye drops (timolol maleate or latanoprost); and (2) the single treatment with SLT.

## 2.0 Methods

For economic feasibility evaluation, as the effectiveness of initial treatment with prostaglandin analog eye drops is like to SLT, cost-minimization analysis (CMA) was used. In CMA, the cost difference between alternative interventions that are assumed to produce equivalent results is calculated, differing only in their incurred costs. When two strategies have the same therapeutic efficacy and the same health outcomes but different costs, the lower-cost strategy is preferable.(11)

An economic impact analysis was conducted according to the recommendations of the Brazilian Ministry of Health using data on the ocular disease glaucoma, which affects individuals over 40 years old. The study population size and characteristics were calculated based on prevalence estimates for the Brazilian population. The efficacy and safety of the two treatments are well established in the literature and are similar.

The assumption was that SUS would have to provide one of the following treatments to the entire target population for 48 months:

1. Latanoprost eye drops
2. Timolol eye drops
3. SLT

### 2.1 Target Population

A hypothetical cohort of 5,000 individuals with primary open-angle glaucoma (POAG) over 40 years old was adopted. Subsequently, to demonstrate the budgetary impact on public funds, this number was projected for the 2021 estimates of the Brazilian population by the Brazilian Institute of Geography and Statistics (IBGE). An average prevalence of POAG of 3,0% of the population over 40 years old was used, based on international epidemiological studies and the national study by Sakata et al.(4)

### 2.2 Medication and Procedure Costs

#### 2.2.1 Medication

The cost of a bottle of latanoprost or timolol eye drops, assuming one unit lasts 30 days when used in both eyes of an individual, was obtained from the maximum consumer price provided by the Drug Market Regulation Chamber. The average acquisition cost was also considered by accessing the Price Panel of the Brazilian Ministry of Economy, given that purchasing through price quotations in larger quantities and at lower costs is mandatory in the federal public service.

It was decided not to adopt the cost of eye drops paid by the Glaucoma Program of the Brazilian Ministry of Health because the program links the price of the medication to the cost of the consultation and some tests.

#### 2.2.2 Laser Procedure

The cost of the SLT procedure is in accordance with the Management System of the SUS Table of Procedures, Medications, and Medical Supplies (SIGTAP). The total cost of two procedures, one for each eye, for the same period, according to efficacy studies, was considered(9). Additionally, expenses (FOB price, free on board, in Miami, USA) for the acquisition (single), installation (single), operation (annual), and maintenance (annual) of the laser equipment, as well as its depreciation, were added to the cost of scenario 3.

The average depreciation rate for equipment is 10% per year with a useful life of 10 years. Depreciation = (AC - RV) x r, where “AC” is the acquisition cost of the asset, “RV” is its residual value, and “r” is the corresponding depreciation rate.

### 2.3 Sensitivity Analysis

The cost difference of the alternative scenario (SLT) was calculated in comparison to two reference scenarios (use of latanoprost eye drops or timolol maleate eye drops) over one and five-year analytical horizons(12,13).

## 3.0 Results

Based on a hypothetical sample of 5,000 individuals and subsequently applying the 3% prevalence of primary open-angle glaucoma in the population over 40 years old to the IBGE population size for 2021, the economic impact on SUS was calculated for the most common initial glaucoma treatment scenarios: (1) with first-choice eye drops (prostaglandin analogs or beta-blockers) or (2) with laser therapy (SLT) over one and five years.

The prices of medications from the latest government purchases (Ministry of Economy Price Panel website: https://paineldeprecos.planejamento.gov.br/ - accessed on 06/29/2022), as well as the maximum consumer price (Drug Market Regulation Chamber - CMED - National Health Surveillance Agency; updated on 06/29/2022), and the SIGTAP (SUS) table for the SLT procedure (code 04.05.05.012) were considered and converted from Brazilian Real (BRL) to United States Dollar (USD) at an exchange rate of 5,2 (Central Bank of Brazil, 06/29/2022). The laser equipment had its FOB price (free on board; in Miami, USA) of approximately USD 43.269,23 considered.

For the first scenario (Table 1), a non-conservative analysis was adopted: the best scenario for eye drops, i.e., constant values from the Ministry of Economy Price Panel and distribution of 70% of individuals using timolol maleate (lower cost) and the rest using latanoprost (higher cost between the two). In this scenario, the economic impact of the treatments is similar over one year and lower for SLT over five years, with a difference of USD 346.961,54 for 5,000 individuals and reaching USD 437.171.538,46 when considering the national population prevalence.

**Table 1.**
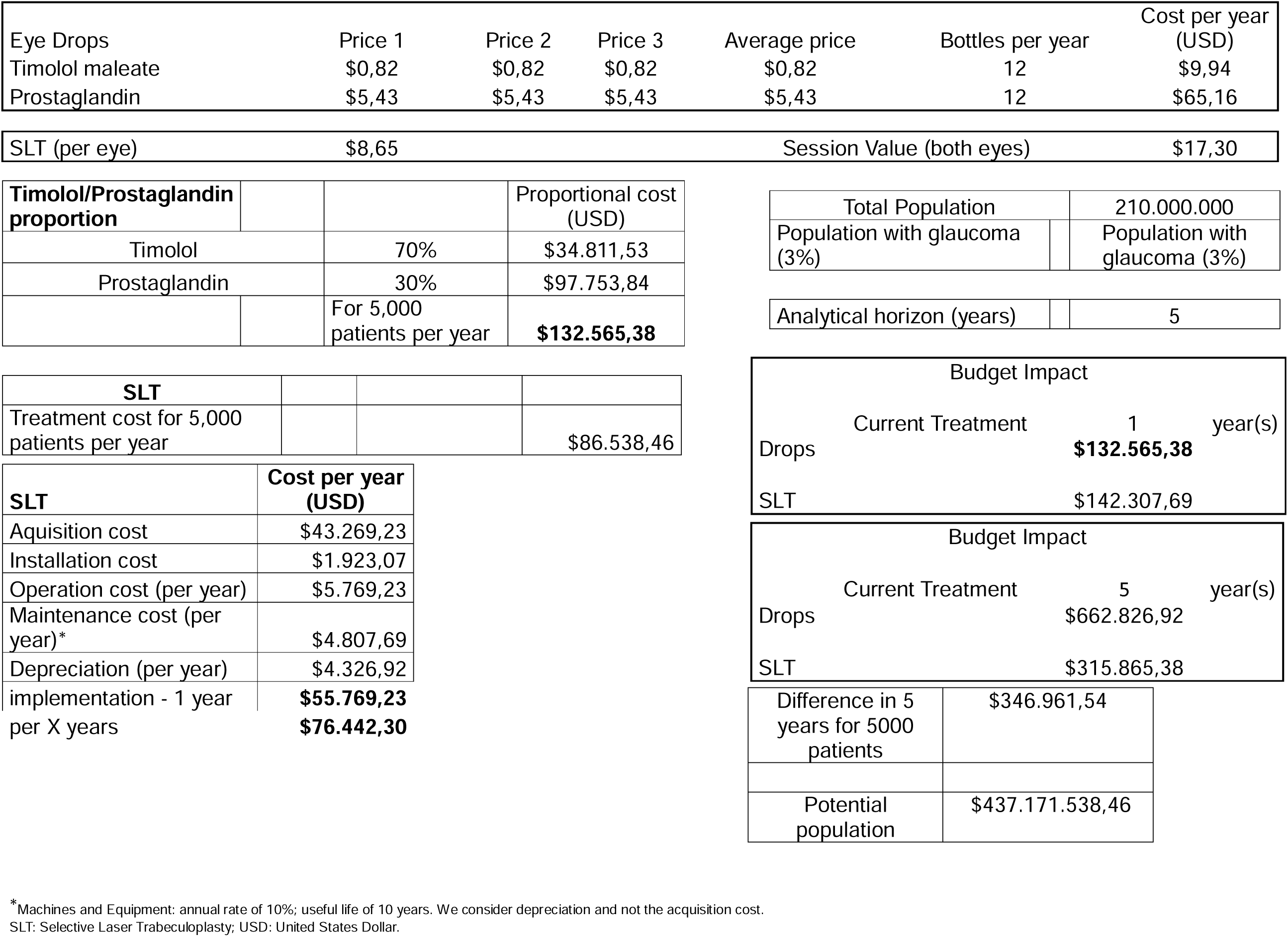
Economic Impact - Non-Conservative Analysis (Best Case Scenario for Eye Drop Type Distribution).

For the second scenario (Table 2), a conservative analysis was adopted: the worst scenario for eye drops, i.e., constant values from the Ministry of Economy Price Panel and distribution of 70% of individuals using latanoprost (higher cost) and the rest using timolol (lower cost between the two). In this scenario, the economic impact of the treatments is already advantageous for SLT over one year (more than half a million reais for 5,000 people) and much lower for laser over five years, with a difference of USD 899.192,30 for 5,000 individuals, projecting to USD 1.132.982.307,69 when considering the disease prevalence in the national population.

**Table 2.**
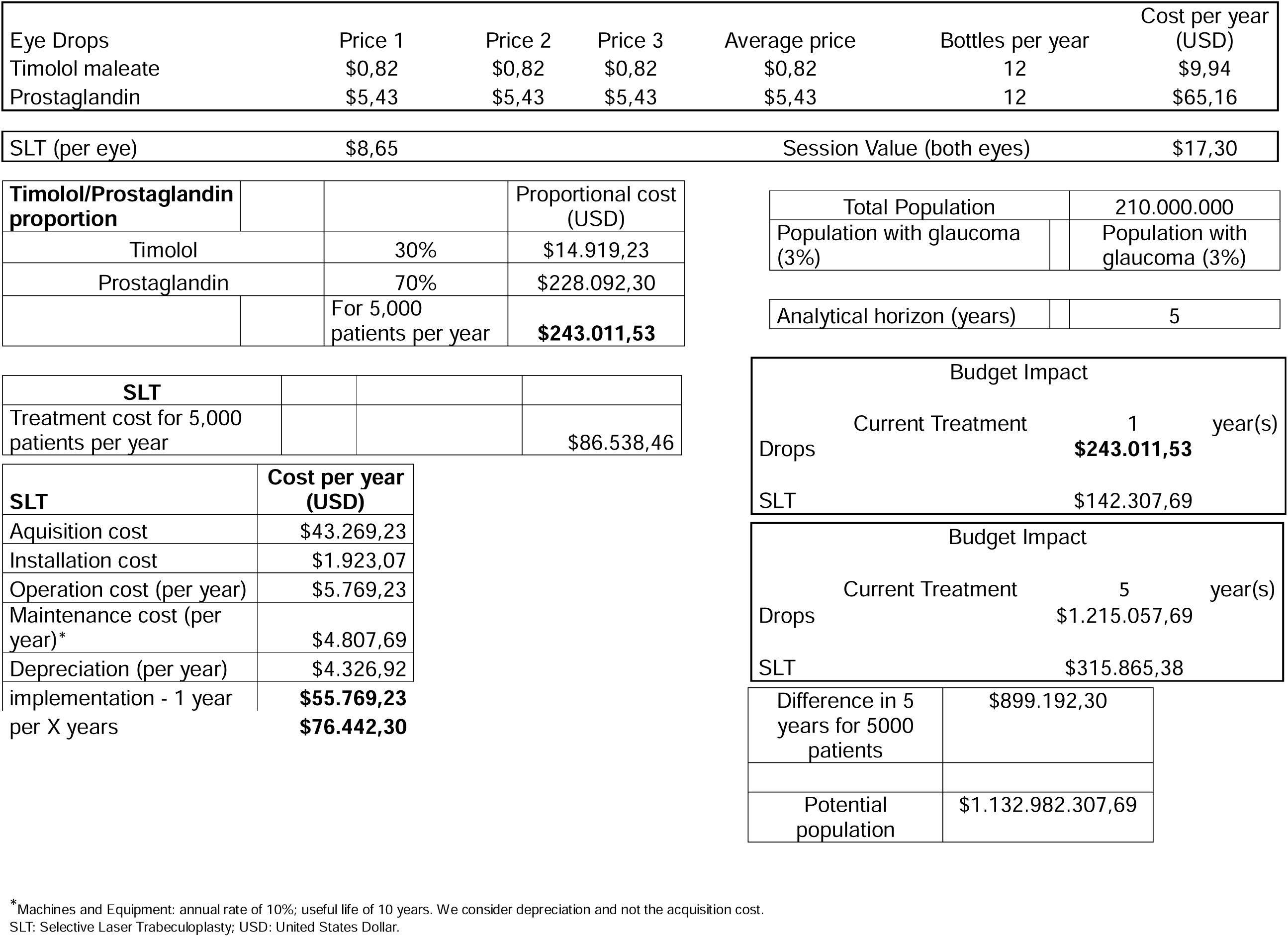
Economic Impact - Conservative Analysis (Worst Case Scenario for Eye Drop Type Distribution)

| Eye Drops | Price 1 | Price 2 | Price 3 | Average price | Bottles per year | Cost per year (USD) |
| --- | --- | --- | --- | --- | --- | --- |
| Timolol maleate | \$0,82 | \$0,82 | \$0,82 | \$0,82 | 12 | \$9,94 |
| Prostaglandin | \$5,43 | \$5,43 | \$5,43 | \$5,43 | 12 | \$65,16 |

|  |  |  |  |
| --- | --- | --- | --- |
| SLT (per eye) | \$8,65 | Session Value (both eyes) | \$17,30 |

| Timolol/Prostaglandin proportion |  | Proportional cost (USD) |
| --- | --- | --- |
| Timolol | 30% | \$14.919,23 |
| Prostaglandin | 70% | \$228.092,30 |
| | For 5,000 patients per year | **\$243.011,53** |

| SLT |  |  |  |
| --- | --- | --- | --- |
| Treatment cost for 5,000 patients per year | | | \$86.538,46 |

|  |  |
| --- | --- |
| Total Population | 210.000.000 |
| Population with glaucoma (3%) | Population with glaucoma (3%) |
| Analytical horizon (years) | 5 |

| Budget Impact |  |  |
| --- | --- | --- |
| Current Treatment | 1 | year(s) |
| Drops | **\$243.011,53** | |
| SLT | \$142.307,69 | |
| Current Treatment | 5 | year(s) |
| Drops | \$1.215.057,69 | |
| SLT | \$315.865,38 | |

|  |  |
| --- | --- |
| Difference in 5 years for 5000 patients | \$899.192,30 |
| Potential population | \$1.132.982.307,69 |

\*Machines and Equipment: annual rate of 10%; useful life of 10 years. We consider depreciation and not the acquisition cost. SLT: Selective Laser Trabeculoplasty; USD: United States Dollar.

Finally, in the third scenario (Table 3), a super conservative analysis was adopted: the worst scenario of eye drop distribution and maximum consumer price, i.e., constant values from the Drug Market Regulation Chamber (the average of the two lowest values and the highest value due to the incidence of the Brazilian Tax on the Circulation of Goods and Services - ICMS) and distribution of 70% of individuals using latanoprost (higher cost) and the rest using timolol (lower cost between the two). In this scenario, the economic impact of the treatments is already extremely advantageous for SLT over one year and much lower for laser over five years, with a difference of USD 6.699.096,15 for 5,000 individuals, reaching a substantial USD 8.440.861.153,84 when considering the national prevalence of glaucoma.

**Table 3.**
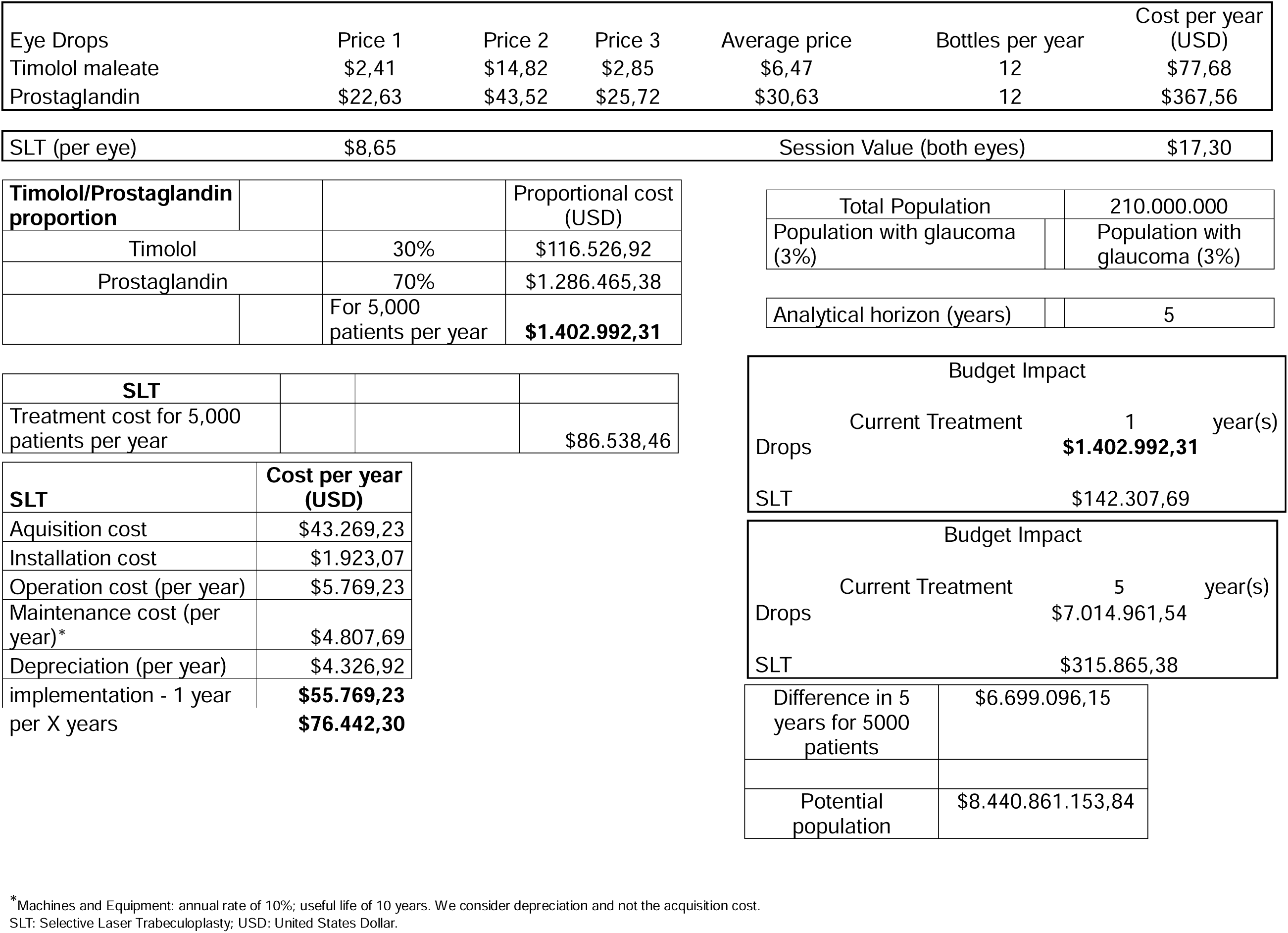
Economic Impact - Conservative Analysis 2 (Worst Case Scenario for Eye Drop Type Distribution + Maximum Consumer Price)

| Eye Drops | Price 1 | Price 2 | Price 3 | Average price | Bottles per year | Cost per year (USD) |
| --- | --- | --- | --- | --- | --- | --- |
| Timolol maleate | \$2,41 | \$14,82 | \$2,85 | \$6,47 | 12 | \$77,68 |
| Prostaglandin | \$22,63 | \$43,52 | \$25,72 | \$30,63 | 12 | \$367,56 |

|  |  |  |  |
| --- | --- | --- | --- |
| SLT (per eye) | \$8,65 | Session Value (both eyes) | \$17,30 |

| Timolol/Prostaglandin proportion |  | Proportional cost (USD) |
| --- | --- | --- |
| Timolol | 30% | \$116.526,92 |
| Prostaglandin | 70% | \$1.286.465,38 |
| | For 5,000 patients per year | **\$1.402.992,31** |

| SLT |  |  |  |
| --- | --- | --- | --- |
| Treatment cost for 5,000 patients per year | | | \$86.538,46 |

|  |  |
| --- | --- |
| Total Population | 210.000.000 |
| Population with glaucoma (3%) | Population with glaucoma (3%) |
| Analytical horizon (years) | 5 |

| Budget Impact |  |  |
| --- | --- | --- |
| Current Treatment | 1 | year(s) |
| Drops | **\$1.402.992,31** | |
| SLT | \$142.307,69 | |
| Current Treatment | 5 | year(s) |
| Drops | \$7.014.961,54 | |
| SLT | \$315.865,38 | |

|  |  |
| --- | --- |
| Difference in 5 years for 5000 patients | \$6.699.096,15 |
| Potential population | \$8.440.861.153,84 |

\*Machines and Equipment: annual rate of 10%; useful life of 10 years. We consider depreciation and not the acquisition cost. SLT: Selective Laser Trabeculoplasty; USD: United States Dollar.

## 4.0 Discussion

The treatment of patients diagnosed with primary open-angle glaucoma (POAG) has traditionally begun with the continuous daily use of hypotensive eye drops. First-line treatments in this therapy, according to medical society protocols and Brazilian Ministry of Health agencies, include beta-blockers (timolol maleate 0.5%) with twice-daily dosing and lower cost, and prostaglandin analogs (latanoprost, bimatoprost, travoprost, and tafluprost) used once at night and with higher cost. Chronic use of eye drops poses challenges and limitations, such as cost and adherence to treatment, potential reduction in quality of life, local and systemic side effects from both the active ingredient and preservatives, among others.

In contrast, SLT is a one-time procedure with a therapeutic result proven to be similar to the continuous use of a first-line hypotensive eye drop. However, it depends on laser equipment, which has acquisition and maintenance costs, as well as depreciation, and requires a trained ophthalmologist to perform it.

The three analyzed scenarios demonstrated a significantly lower economic impact of SLT for SUS over one to five years, with a favorable difference for laser of more than USD 8 billion over five years when considering 3% of the Brazilian population in 2021. The public manager would have unquestionable economic advantage if adopting SLT as the initial therapy for POAG in Brazil, even allowing for the improvement of reimbursements for the procedure, which still includes an old class of laser (argon) when the selective YAG laser is already used; hence the current nomenclature of selective laser trabeculoplasty (SLT).

## 5.0 Conclusion

The economic impact on the Brazilian Public Health System (SUS) was lower for Selective Laser Trabeculoplasty (SLT) compared to the use of latanoprost and timolol maleate eye drops as the initial treatment for primary open-angle glaucoma in all scenarios studied over one and five-year periods.

## Data Availability

All data produced in the present work are contained in the manuscript

## 6.0 Author Contributions

Significant contribution to conception and design: Ivan M. Tavares; Flavio E. Hirai; Diogo F. C. Landim, Paola Zucchi

Data Acquisition: Ivan M. Tavares, Flavio E. Hirai

Data Analysis and Interpretation: Ivan M. Tavares; Flavio E. Hirai; Paola Zucchi

Manuscript Drafting: Ivan M. Tavares; Diogo F. C. Landim; Flavio E. Hirai, Paola Zucchi

Significant intellectual content revision of the manuscript: Ivan M. Tavares, Diogo F. C. Landim, Flavio E. Hirai; Paola Zucchi

Have given final approval of the submitted manuscript (mandatory participation for all authors): Ivan M. Tavares; Flavio E. Hirai; Diogo F. C. Landim; Paola Zucchi

Statistical analysis: Flavio E. Hirai

Obtaining funding: Not applicable.

Supervision of administrative, technical, or material support: Ivan M. Tavares, Flavio E. Hirai; Diogo F. C. Landim; Paola Zucchi

Research group leadership: Flavio E. Hirai, Paola Zucchi, Ivan M Tavares

